# Unusual patterns of risk for diabetic retinopathy in the Mumbai slums: The Aditya Jyot Diabetic Retinopathy in Urban Mumbai Slums Study (AJ-DRUMSS) Report 3

**DOI:** 10.1101/2022.03.16.22272514

**Authors:** Radhika Krishnan, Astha Jain, Siddhita Nare, Rajkumar Shankar, Jacquelaine Bartlett, Sudha K. Iyengar, Scott M. Williams, Natarajan Sundaram

**Affiliations:** Aditya Jyot Foundation for Twinkling Little Eyes, Mumbai, Maharashtra, India; Department of Population and Quantitative Health Sciences, Case Western Reserve University, Cleveland, OH, USA

**Keywords:** Diabetic retinopathy, Epidemiology, Slum population, Obesity

## Abstract

Diabetes onset precedes diabetic retinopathy (DR) by 5-10 years, but many people with diabetes remain free of this microvascular complication. Our aim was to identify risk factors for DR progression in a unique and diverse population, the slums of Mumbai.

We performed a population-based cross-sectional analysis of 1163 diabetics over 40 years of age from slums in 18 wards of Mumbai. Data was collected on 33 variables and assessed for association with DR using both univariate and multivariate analyses. Stratified analyses were also performed on males and females, separately. Among hypertensive individuals we also assessed whether duration of hypertension associated with DR.

Of 31 non-correlated variables analysed as risk factors for DR, 15 showed evidence of significant association. The most prominent included sex, where being a female associated with decreased risk of DR, while longer duration of diabetes and poor glycaemic control associated with increased risk. The duration of diabetes risk was partially, but significantly, mediated by age of diabetes diagnoses (8.6% of variance explained, p = 0.012). Obesity as measured by several measures, including body mass index (BMI) and measures of central obesity had a negative association with DR; increased measures of obesity consistently reduced risk of DR.

There was some agreement with risk factors described in earlier studies (e.g., duration of diabetes and glycaemic control), but other factors such as obesity measures appeared to have a reversed direction of effect compared to most prior studies. These results indicated that the overall pattern of association in the Mumbai slums was novel. Thus, in previously uncharacterized populations, such as the slums that we examined, it is important to evaluate all risk factors *de novo* to appropriately assess patterns of risk.

## Introduction

Almost 20% of the world’s population resides in India, and this South Asian country endures an exceptional burden of diabetes mellitus (DM) with ∼70 million people at present and a predicted 100 million by 2030 (1-4). Given this epidemic of DM, diabetic retinopathy (DR) presents a huge burden of avoidable blindness(5), especially since Indians are more likely to develop both DM(6) and DR (7, 8). Unfortunately, both DM and DR are frequently undetected until irreparable vision loss occurs, making it critical to understand key associating risk factors, especially in high-risk settings that can allow early intervention.

DR risk is greatest in people with uncontrolled DM, as extended hyperglycemia causes damage to retinal vasculature, frequently partitioned into, non-proliferative diabetic retinopathy (NPDR) and proliferative diabetic retinopathy (PDR), with or without diabetic macular edema (DME). In India >50% of people with diabetes have poor glycemic control(9, 10), uncontrolled hypertension and dyslipidemia(11). There is reason to suspect that DR risks are novel to India as Indians have a high prevalence of low body mass index (BMI) DM and a young age of onset(12). And within India, the urban poor, i.e., slum dwellers, are more likely to suffer diabetic complications, especially DR(13). In the slums, inadequate DM treatment results in poor DM control and lack of adherence to science-based modern medications, making slum dwellers more vulnerable to DR. This population may provide additional insights important to blindness reduction due to unique patterns of risk and novel combination of exposures.

In East and South Asia, higher HbA1c, duration of diabetes, male gender, microalbuminuria and insulin therapy are risk factors of DR (14) (15). Other associated risk factors include anemia(16), lipid abnormalities(17), systemic hypertension(18, 19), pregnancy(20, 21), nephropathy(22) and cardiovascular disease (CVD)(23). However, the prevalence of these risk factors may be different in Indian slums due to variation in risk factor exposures (24). Hence, it is important to assess risk factors that are or may be different for DM than those in Europe(4, 25), as variability in the presentation of DM factors may affect DR risk.

In this study, we sought to understand how risk factors, common in slum settings, associate with DR and modify each other. We used our already developed research and clinical infrastructure to perform studies of DR in the high-risk Mumbai slums (26). Although the proportion of NPDR observed was comparable to that observed elsewhere in Southern India(27), PDR was nearly three times higher than other Indian populations, and sight-threatening DR (STDR) almost two-fold higher than rural India(28). These patterns are comparable to other migrant Indian populations across Asia(29). Our goal was to assess associations in DR among slum dwellers and to see the extent to which this unique population presents with different or similar risks to other studied populations. Our study emphasized known and unknown clinical factors in a high-risk population that could easily be assessed in low resource settings, as early treatment is critical to preventing blindness.

## Methods

The study protocol was approved by the Institutional Ethics Committee of Aditya Jyot Eye Hospital, Mumbai, India AJEHEC2016/21/01 and adhered to the tenets of the Declaration of Helsinki [https://www.wma.net/what-we-do/medical-ethics/declaration-of-helsinki/]. Written informed consent was obtained from the study participants and all privacy requirements were met.

### Recruitment of subjects and collection of data

Study participants consisted of 1163 diabetic subjects ≥ 40 years of age who were recruited as part of our previous population-based cross-sectional prospective study conducted in Mumbai (26, 27). Subjects were recruited from 18 municipal wards that included slum populations. Diagnosis of diabetes mellitus (DM) was made using one of the following criteria: 1) prior diagnosis by medical practitioner or 2) fasting plasma glucose was greater than or equal to 126 mg/dL measured by glucometer (Ascensia Entrust, Bayer Diagnostics, Tarrytown, NY, USA), as per the guidelines of American Diabetic Association. Subjects were categorised into known diabetic (KD), provisional diabetic (PD), or not known diabetic (NKD)(26). People categorized as NKD underwent preliminary visual acuity examinations but were not included in the final cohort, while those categorised as PD or KD underwent DR screening. Only individuals with confirmed diabetes were included in our analyses. Data on an additional 33 variables were also collected for all study participants (18 Dichotomized, 2 Categorical and 13 Continuous variables in all) (**Table S1**).

Variables were selected based on prior studies indicating putative association. The thirty-three exposure variables were determined, and descriptive statistics were computed for all the variables (see **Table 1**). The exposure variables included: sex, age, hypertension, smoking, ear lobe crease, polyuria, polydipsia, weight loss, ischemic heart disease, stroke, neuropathy, nephropathy, family history of diabetes, vegetarian diet, central obesity score (coded based on waist to hip ratio (WHR) with Female subjects WHR <0.85 and Male subjects WHR <0.95 considered as normal, whereas Female subjects with WHR ≥ 0.85 and Male subjects WHR ≥ 0.95 as obese), abdominal circumference code (abdominal circumference coding was done as follows: abdominal circumference of male subjects ≤102cm and female subjects ≤88cm considered as normal, whereas abdominal circumference of male subjects >102cm; female subjects >88cm considered as obese), literacy, religion, diabetes treatment, occupation, food, fasting plasma glucose, systolic blood pressure, diastolic blood pressure, duration of diabetes, weight, height, body mass index, waist to hip ratio, abdominal circumference, hip circumference, abdominal to hip ratio, and duration of diabetes treatment.

**Table 1.** Variables included in analyses.

| **Variables** | **Total (n=1163)** | **No DR (n=905)** | **DR (n=258)** |
| --- | --- | --- | --- |
| **A. DICHOTOMOUS VARIABLES (NUMBER (%))** |  |  |  |
| **SEX** |  |  |  |
| Women | 618 (53.1) | 509 (56.2) | 109 (42.2) |
| Men | 545 (46.9) | 396 (43.8) | 149 (57.8) |
| **HYPERTENSION** |  |  |  |
| Yes | 609 (52.5) | 466 (51.7) | 143 (55.4) |
| No | 551 (47.5) | 436 (48.3) | 115 (44.6) |
| **SMOKING** |  |  |  |
| Ever | 236 (20.3) | 166 (18.4) | 70 (27.1) |
| Never | 926 (79.7) | 738 (81.6) | 188 (72.9) |
| **EAR LOBE CREASE** |  |  |  |
| Present | 88 (8.4) | 58 (7.1) | 30 (12.9) |
| Absent | 966 (91.7) | 763 (92.9) | 203 (87.1) |
| **POLYURIA** |  |  |  |
| Yes | 849 (73.0) | 642 (70.9) | 207 (80.2) |
| No | 314 (27.0) | 263 (29.1) | 51 (19.8) |
| **POLYDIPSIA** |  |  |  |
| Yes | 838 (72.1) | 633 (69.9) | 205 (79.5) |
| No | 325 (27.9) | 272 (30.1) | 53 (20.5) |
| **WEIGHT LOSS** |  |  |  |
| Yes | 318 (27.3) | 236 (26.1) | 82 (31.8) |
| No | 845 (72.7) | 669 (73.1) | 176 (68.2) |
| **ISCHEMIC HEART DISEASE** |  |  |  |
| Yes | 105 (9.0) | 83 (9.2) | 22 (8.5) |
| No | 1057 (91.0) | 821 (90.8) | 236 (91.5) |
| **STROKE** |  |  |  |
| Yes | 29 (2.5) | 18 (2.0) | 11 (4.3) |
| No | 1132 (97.5) | 885 (98.0) | 247 (95.7) |
| **NEUROPATHY** |  |  |  |
| Yes | 37 (3.2) | 24 (2.7) | 13 (5.0) |
| No | 1125 (96.8) | 880 (97.4) | 245 (95.0) |
| **NEPHROPATHY** |  |  |  |
| Yes | 23 (2.0) | 15 (1.7) | 8 (3.1) |
| No | 1138 (98.0) | 889 (98.3) | 249 (96.9) |
| **FAMILY HISTORY OF DIABETES** |  |  |  |
| Yes | 446 (38.9) | 345 (38.7) | 101 (38.9) |
| No | 700 (61.1) | 546 (61.3) | 154 (60.4) |
| **VEGETARIAN DIET** |  |  |  |
| Non-vegetarian | 971 (90.0) | 753 (89.8) | 218 (90.8) |
| Vegetarian | 108 (10.0) | 86 (10.3) | 22 (9.2) |
| **CENTRAL OBESITY SCORE** |  |  |  |
| Normal | 200 (18.9) | 150 (18.1) | 50 (21.4) |
| Central Obesity | 861 (81.2) | 677 81.9) | 184 (78.6) |
| **ABDOMINAL CIRCUMFERENCE CODE** |  |  |  |
| Normal | 572 (49.2) | 409 (45.2) | 163 (63.4) |
| Obese | 590 (50.8) | 496 (54.8) | 94 (36.6) |
| **LITERATE** |  |  |  |
| Yes | 836 (71.9) | 638 (70.5) | 198 (77.0) |
| No | 326 (28.0) | 267 (29.5) | 59 (23.0) |
| **RELIGION** |  |  |  |
| Muslim | 137 (12.0) | 104 (11.7) | 33 (12.8) |
| Hindu | 1009 (88.1) | 785 (88.3) | 224 (87.2) |
| **TREATMENT** |  |  |  |
| Metformin | 769 (69.9) | 595 (69.6) | 174 (70.7) |
| No metformin | 332 (30.2) | 260 (30.4) | 72 (29.3) |
| **B. CATEGORICAL VARIABLES (NUMBER (%))** |  |  |  |
| **OCCUPATION** |  |  |  |
| Not working | 89 (7.7) | 62 (6.9) | 27 (10.5) |
| Working | 453 (39.0) | 341 (37.7) | 112 (43.6) |
| Housewife | 531 (45.7) | 434 (48.0) | 97 (37.7) |
| Retired | 89 (7.7) | 68 (7.5) | 21 (8.2) |
| **FOOD** |  |  |  |
| Rice only | 27 (2.5) | 22 (2.7) | 5 (2.1) |
| Wheat only | 227 (21.4) | 166 (20.1) | 61 (26.0) |
| Both rice & wheat | 807 (76.1) | 638 (77.9) | 169 (71.9) |
| **C. CONTINUOUS VARIABLES (MEAN ± SD)** |  |  |  |
| Age | 55.60 ± 9.30 | 55.59 ± 9.39 | 55.66 ± 9.01 |
| Fasting Plasma Glucose | 188.61 ± 81.56 | 184.38 ± 78.52 | 203.04 ± 89.89 |
| Systolic Blood Pressure | 139.31 ± 22.44 | 138.33 ± 22.16 | 142.74 ± 23.10 |
| Diastolic Blood Pressure | 80.72 ± 12.17 | 80.74 ± 12.14 | 80.64 ± 12.30 |
| Duration of diabetes | 5.53 ± 5.55 | 4.60 ± 4.82 | 8.80 ± 6.61 |
| Weight | 63.29 ± 12.30 | 63.66 ± 12.65 | 61.99 ± 10.89 |
| Height | 155.75 ± 10.80 | 155.42 ± 10.77 | 156.93 ± 10.86 |
| BMI | 26.25 ± 5.54 | 26.50 ± 5.67 | 25.36 ± 4.96 |
| Waist/Hip Ratio CO | 0.97 ± 0.11 | 0.97 ± 0.12 | 0.96 ± 0.08 |
| Abdominal Circumference | 96.40 ± 13.24 | 97.01 ± 13.53 | 94.24 ± 11.96 |
| Hip circumference | 99.56 ± 11.08 | 99.81 ± 10.98 | 98.68 ± 11.43 |
| Abdominal to Hip Ratio* | 0.97 ± 0.12 | 0.97 ± 0.12 | 0.96 ± 0.09 |
| Duration of Diabetes treatment* | 5.25 ± 5.27 | 4.55 ± 4.70 | 7.48 ± 6.30 |

\*Removed from analyses due to high correlation with other variables (see Methods)

Data were obtained using a validated questionnaire, based on each subject’s self-report or by physical and clinical evaluation (26, 27). Community health workers (CHWs) ensured that the entire questionnaire was filled out completely and accurately. The data in the medical history included duration and treatment of chronic diseases such as diabetes and hypertension, a family history of diabetes, the subject’s degree of relatedness to diabetic family members, symptoms related to complications of diabetes (e.g., diabetic nephropathy and diabetic neuropathy), tobacco intake and alcohol consumption - their form of intake, duration, amount, and age at start, present and past status. The questionnaire and medical history were recorded by the CHWs. All collected data was cleaned and stored in a centralized data storage system. All computers were password protected and protected by a firewall. External hard drives for storage were used for the long-term archival of the data. All recorded data was regularly backed-up on the password protected external hard disk.

A trained CHW measured the blood pressure (BP) of registered subjects with a mercury column sphygmomanometer (Deluxe, BPMR 120, Maharashtra, India) and a stethoscope in seated position after a resting period of 5 minutes. All BP measurements were made on the right arm of each subject, using a cuff of appropriate size at the level of the heart. Two readings were taken 5 minutes apart and the mean of the two was taken as the BP. Hypertension was recorded if the systolic blood pressure was greater than 140 mmHg or the diastolic blood pressure greater than 90mmHg or the individual was on antihypertensive medication. Duration of DM was recorded as time interval between diagnosis of diabetes (as made by a diabetologist or when anti-diabetic treatment was started) and the date of administration of questionnaire.

Height and weight of each subject were measured after which the BMI was calculated as: Weight (kg)/Height (m^2^). Waist circumference was measured 2.5 cm above the umbilicus. Hip circumference was measured at the greatest circumference around the buttocks.

### Ophthalmic Examination and diagnosis of diabetic retinopathy

All diabetic subjects (n=1163) underwent a comprehensive ophthalmic examination, including visual acuity assessment, intraocular pressure (IOP) measurement, anterior segment and dilated posterior segment evaluation (tropicamide 0.5% & phenylephrine 5%). Seven-field 30° stereoscopic paired fundus photographs were obtained using a fundus camera (VISUCAM 500, Carl Zeiss Meditech AG, Jena, Germany), and were graded in an un-masked manner by a single experienced retina specialist (AJ) by the Early Treatment of Diabetic Retinopathy Study (ETDRS) classification system (30, 31). Re-grading photographs of every tenth subject resulted in intra-observer variations within acceptable limits (k=1.0) and served as internal validation for these measures.

Photographic grading of diabetic retinopathy: A 30° stereoscopic pair of color photographs of seven standard fields were captured by using the fundus camera (VISUCAM 500, Carl Zeiss meditech AG, Jena, Germany). Images were stored as uncompressed jpeg files without any enhancements. The grading of DR was based on photographs graded against the standard photographs of the ETDRS done by a single experienced retinal specialist in an unmasked manner to check for any major discrepancies with the clinical grading.

The fundus photographs identified 258 diabetic subjects as having diabetic retinopathy (DR). Although we were mainly concerned with DR among those with Type II diabetes (T2DM), there may have been rare cases of Type 1 diabetes (T1DM) in our study. Given the low prevalence of T1DM in India (32), and as we only enrolled individuals > 40 years of age, the number of T1DM subjects should have been minimal.

### Statistical Analyses

Our outcome of interest was DR among individuals with DM. Prior to a comprehensive regression analysis, we tested for correlation between independent variables using Pearson’s Correlation to ensure that only independent terms were included in the model. This analyses indicated that 31 of the 33 variables tested for correlations were sufficiently independent (absolute value of correlation < 0.8)(**Table S2**). Duration of diabetes and duration of diabetes treatment had a corelation of 0.88 and duration of treatment was removed from subsequent analyses. Abdomen to hip ratio was highly correlated with waist hip ratio – central obesity (WHRCO) with a correlation of 0.97 and abdomen to hip ratio was removed. Sex and occupation had a correlation coefficient of -0.80, but both were kept in the analysis. Also, all continuous variables were tested for linear relationships between the logit and exposure of independent risk factors before proceeding with regression analyses using Box-Tidwell tests(33). None of the variables deviated significantly from linearity (**Table S3**). Post-data cleaning and assessment of correlation, 31 variables were kept for subsequent analysis; variables being removed are indicated by a * in Table 1.

To test which variables best predicted DR status, we performed univariate regression analyses for the 31 variables. Variables were assessed for missingness and as none of the variables had a missingness of greater than 10%, all were included in subsequent analyses (**Table S4**). We set a significance threshold at p < 0.05.

We next performed stepwise regression with all 31 variables to select those that explain DR status; we used an entry and exit criterion of p < 0.15 that resulted in nine explanatory variables. We then ran a full multivariate logistic regression analysis with the nine selected variables for DR as the outcome, including all samples that had no missing data (n = 1075). We considered those with a p value less than 0.05 to be associated with DR.

We also did two stratified analyses. First, we performed analyses on males and females separately to determine if there are different patterns of association in the two sexes. We did this as it is well established that DR is more prevalent in males(34). Second, we performed analyses in people with a diagnosis of hypertension separately (n = 593) as there is evidence for association with DR(35). In addition to association with the other measured variables, among those with hypertension we tested for association of duration of hypertension with DR. All analysis was run using SAS/Base®9.4(TS1M6) v15.1 software (SAS Institute Inc.) [35, Cary NC, 2016].

## Results

The demographic and baseline characteristics of 1163 subjects with and without DR are shown in **Table 1**. Despite the majority of participants being female (53.1%), the majority of DR cases were male (57.8%). Age was similar between cases and controls (mean 55.6 in both). Missingness was limited for all variables to under 10%.

Univariate regression analyses comparing cases with DR (n=258) to those without DR (n=905) showed a total of 15 significant associations of 31 comparisons. Being female was associated with reduced risk of DR (OR = 0.57, p < 0.0001). In contrast, being a smoker, having an ear lobe crease, having polyuria, polydipsia, or a history of stroke was associated with increased risk of DR. Obesity as measured by abdominal circumference code appeared to be protective (OR = 0.48, p < 0.0001), while being literate associated with increased risk (OR = 1.40, p= 0.040) (**Table 2A**). Presence of neuropathy was borderline significant with p = 0.059 (**Table S5**). Of the two categorical variables one, occupation, showed evidence of association, as being a housewife associated with reduced risk of DR compared to working outside of the house (OR = 0.68, p = 0.005)(**Table 2B**). Six of eleven continuous variables associated with DR in our study population. These were fasting plasma glucose, systolic blood pressure, duration of DM, height, BMI, and abdominal circumference. Age did not associate with DR, but weight was close to significant (p= 0.055)(**Table S5**). DR associated with higher fasting plasma glucose, higher systolic blood pressure, longer duration of DM, while BMI and abdominal circumference had inverse relationships with DR. All results are presented in **Table S5**.

**Table 2.**
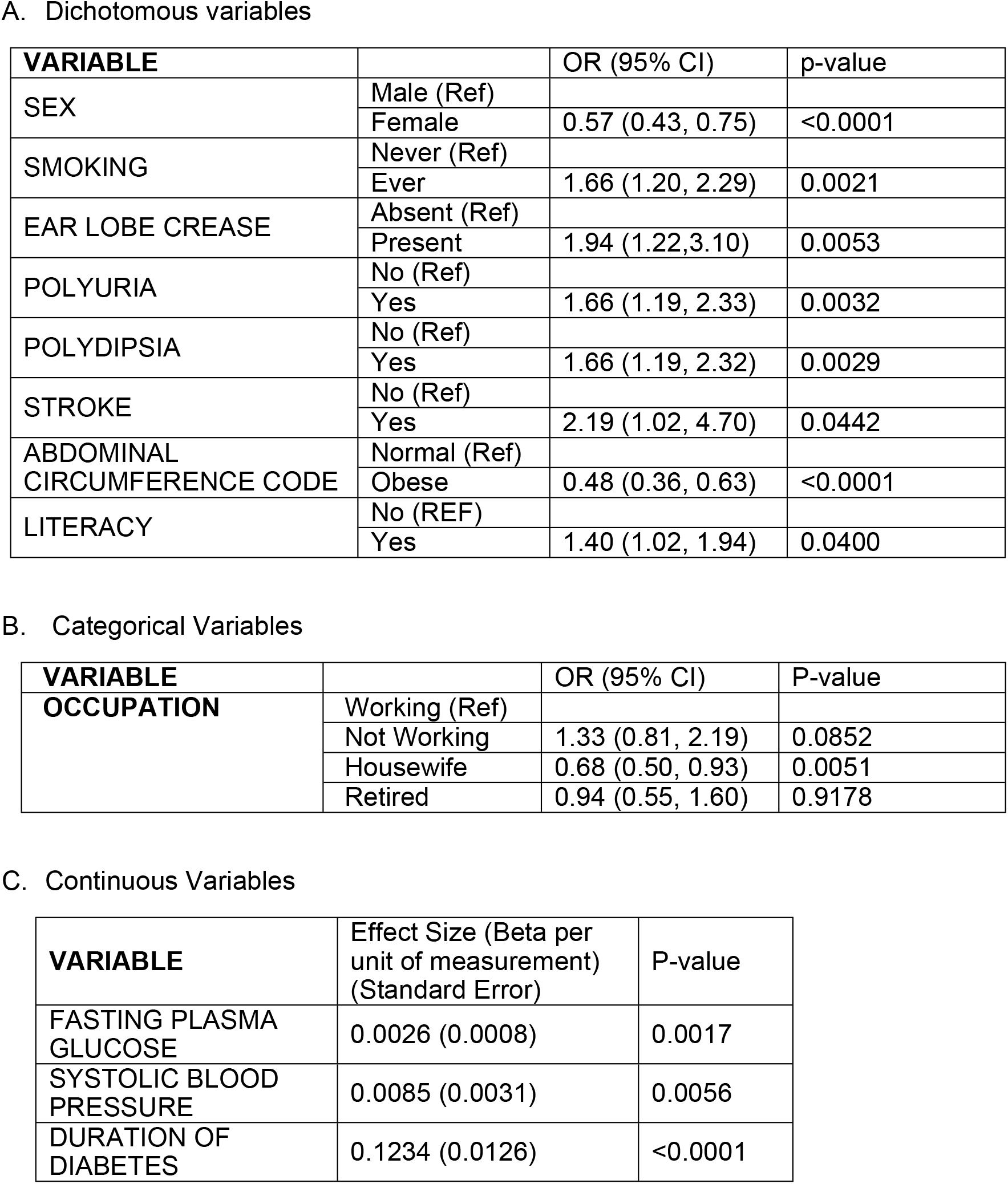
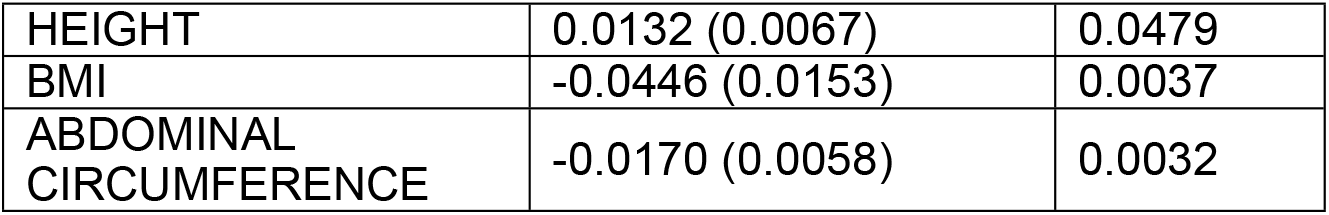
Significant variables from univariate analyses.

Stepwise multivariate regression analyses were performed using all variables. Of the original 31 variables nine met our threshold in stepwise analyses (p < 0.15). The 9 variables and their significance in the stepwise analyses were having neuropathy (p = 0.05), a family history of DM (p = 0.02), abdominal circumference code (p < 0.0001), vegetarian diet (p = 0.146), age (p = 0.0006), fasting plasma glucose (p = 0.0012), both systolic (p = 0.0008) and diastolic blood pressure (p = 0.092), and duration of DM (p < 0.0001). We then ran logistic regression analysis with only these nine variables and seven were significant at p < 0.05 (**Table 3**). The final model indicated that having neuropathy increased risk (OR = 2.33) while having increased central obesity or a family history of DM decreased risk (OR = 0.49 and OR = 0.65, respectively). For continuous variables, age had an inverse relationship with DR risk as did diastolic blood pressure, but higher systolic blood pressure, higher FPG, and longer duration of DM all associated with increased risk (**Table 3**).

**Table 3.** Multivariate analysis using only variables selected empirically by stepwise regression.

| Variable | Reference | Odds Ratio | 95% CI | P value |
| --- | --- | --- | --- | --- |
| **Dichotomous Variables** |  |  |  |  |
| Neuropathy | No | 2.33 | 1.00,5.41 | 0.0500 |
| Family History of DM | No | 0.65 | 0.46,0.94 | 0.0200 |
| Abdominal Circumference Code | Normal | 0.49 | 0.34,0.69 | <0.0001 |
| Vegetarian Diet | Non-Vegetarian | 0.65 | 0.94 | 0.1459 |
| **Continuous Variables** |  |  |  |  |
|  |  | Beta | SE |  |
| Age |  | -0.0385 | 0.0112 | 0.0006 |
| Fasting plasma glucose |  | 0.00327 | 0.001 | 0.0012 |
| Systolic BP |  | 0.0173 | 0.0048 | 0.0008 |
| Diastolic BP |  | -0.0167 | 0.00889 | 0.0918 |
| Duration of DM |  | 0.1360 | 0.0149 | < 0.0001 |

### Subset analyses Sex

Since sex associated with DR, we performed sex stratified analyses to determine whether the associations showed evidence of different patterns of association in males and females. In males, univariate analyses indicated six of the 31 variables associated with DR: neuropathy, abdominal circumference code, fasting plasma glucose, duration of DM, weight and abdominal circumference. Of these, neuropathy associated with increased risk of DR as did duration of DM and fasting plasma glucose (**Table 4**). There were also six associations in females. Polyuria, polydipsia, and stroke all associated with increased risk of DR, whereas obesity as measured by abdominal circumference code associated with decreased risk. Higher systolic blood pressure and longer duration of DM also associated with increased risk (**Table 4**).

**Table 4.**
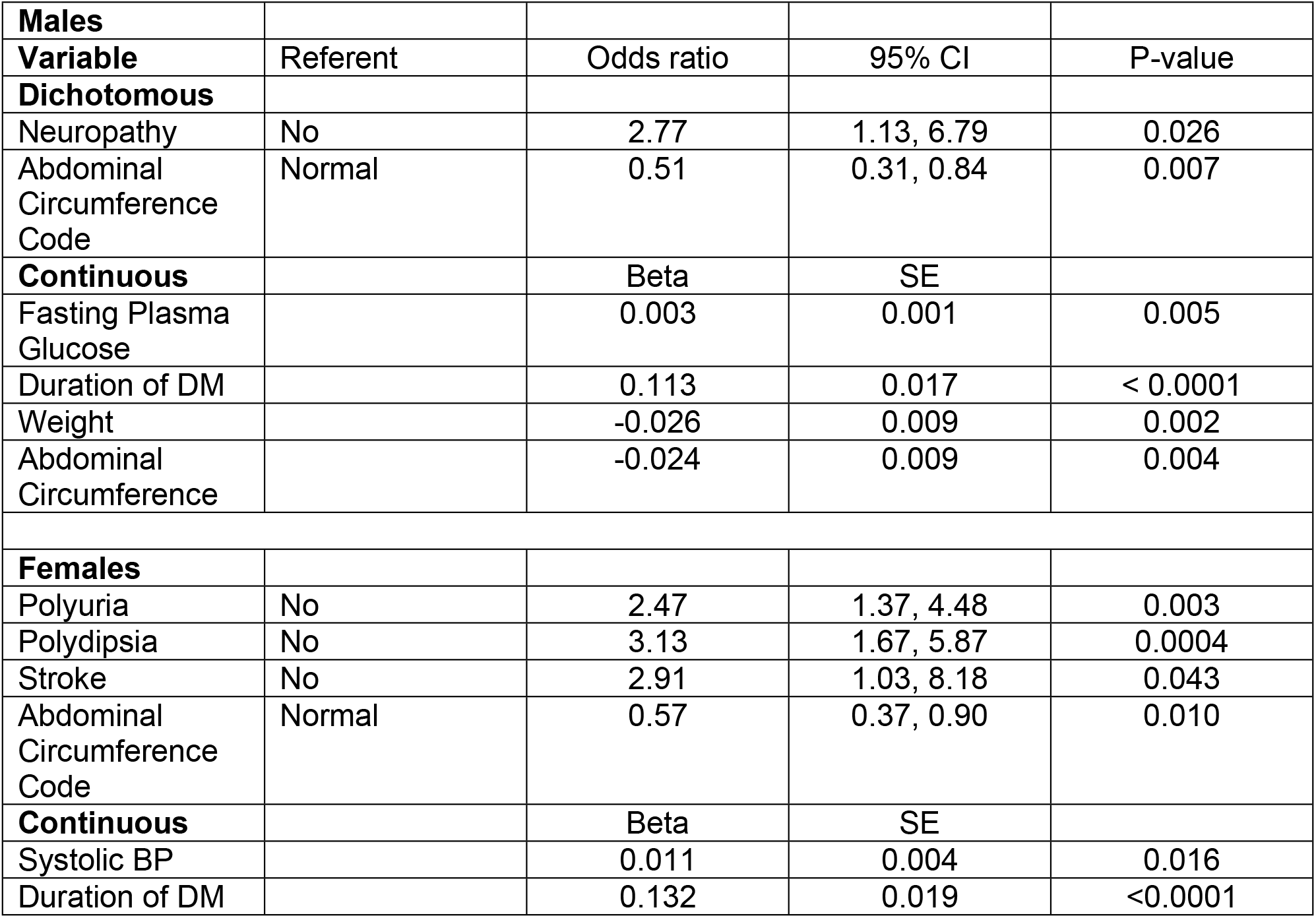
Sex stratified analyses – univariate analyses.

In males, stepwise regression selected 12 variables for the final model. They were ear lobe crease, ischemic heart disease, stroke, neuropathy, family history of DM, vegetarian diet, abdominal circumference code, age, systolic blood pressure, fasting plasma glucose, duration of DM, and weight. Of these ischemic heart disease, neuropathy, age, duration of DM, fasting plasma glucose, and weight were significant in the model that included all 12 variables (**Table 5A, 5B**). Having ischemic heart disease associated with decreased risk of DR. Systolic blood pressure, duration of DM and fasting plasma glucose had positive betas, while age and weight had negative beta values (**Table 5B**).

**Tale 5.** Stepwise multivariate Sex stratified analysis – Males.

**Tale 5. Stepwise multivariate Sex stratified analysis – Males****5A. Dichotomous Variables - Males**
| **Variable** | **Referent** | **OR** | **95% CI** | **P-value** |
| --- | --- | --- | --- | --- |
| Ear Lobe Crease | Absent | 1.86 | 0.86,4.02 | 0.112 |
| Ischemic Heart Disease | No | 0.24 | 0.07, 0.84 | 0.025 |
| Stroke | No | 5.47 | 0.75, 39.94 | 0.094 |
| Neuropathy | No | 7.05 | 2.11, 23.51 | 0.0015 |
| Family History of DM | No | 0.68 | 0.41, 1.10 | 0.116 |
| Vegetarian Diet | Non-Vegetarian | 0.39 | 0.14, 1.09 | 0.073 |
| Abdominal Circumference Code | Normal | 0.62 | 0.33, 1.20 | 0.156 |

**5B. Continuous Variables - Males**
| **Variable** | **Beta** | **SE** | **P-value** |
| --- | --- | --- | --- |
| Age | -0.055 | 0.015 | 0.0003 |
| Systolic BP | 0.010 | 0.005 | 0.057 |
| Duration of DM | 0.1513 | 0.022 | < 0.0001 |
| Fasting Plasma Glucose | 0.0037 | 0.0014 | 0.007 |
| Weight | -0.0266 | 0.012 | 0.030 |

In females, stepwise regression selected eight variables: polydipsia, rice versus wheat eaters, age, systolic and diastolic blood pressure, duration of DM, BMI, and Waste to hip ratio – central obesity. Five of these significantly associated with DR in the model that included all eight variables. Of these polydipsia, systolic blood pressure, and duration of DM positively associated positively with DR, while age and diastolic blood pressure negatively associated with DR (**Table 6**).

**Table 6.** Stepwise multivariate Sex stratified analysis – Females.

6A. Dichotomous and categorical variables
| Variable | Referent | OR | 95% CI | P-value |
| --- | --- | --- | --- | --- |
| Polydipsia | No | 2.26 | 1.10, 4.62 | 0.0260 |
| Rice and wheat eaters | Rice only |  |  |  |
| Both rice and wheat eaters |  | 1.26 | 0.24,6.63 | 0.6023 |
| Wheat only |  | 2.55 | 0.46, 14.17 | 0.0960 |

6B. Continuous Variables
| Variable | Beta | SE | P-value |
| --- | --- | --- | --- |
| Age | -0.035 | 0.016 | 0.028 |
| Systolic BP | 0.025 | 0.007 | 0.0004 |
| Diastolic Blood Pressure | -0.041 | 0.014 | 0.002 |
| Duration of DM | 0.134 | 0.023 | < 0.0001 |
| BMI | -0.049 | 0.027 | 0.067 |
| Waist to hip ratio – central obesity | -1.19 | 1.13 | 0.295 |

### Hypertension

Lastly, as DR associated with systolic blood pressure in most univariate and multivariate analyses, we assessed whether among those with hypertension, if duration of hypertension associated with DR, and it did in a univariate analysis (beta = 0.0675 per year of hypertension; p = 0.0002). Univariate analyses within the hypertensive only group indicated that ten variables associated with DR: sex, ear lobe crease, polyuria, abdominal circumference code, occupation, age, duration of DM, fasting plasma glucose, abdominal circumference, and duration of hypertension (**Table 7**). Being female appeared to reduce risk as did having an obese abdominal circumference code, whereas having an ear lobe crease or polyuria increased risk. Compared to working outside of the house being a housewife reduced risk but being unemployed increased risk. Increasing age, fasting plasma glucose, and duration of DM increased risk. The direction of effects was consistent with prior analyses except age (**Table 7**).

**Table 7.** Univariate association analyses among those with a hypertension diagnosis (n= 609)

A. Categorical Variables
| Variable | Referent | Odds Ratio | 95% CI | P-value |
| --- | --- | --- | --- | --- |
| Sex | Male | 0.55 | 0.38, 0.80 | 0.002 |
| Ear Lobe Crease | Absent | 2.07 | 1.20, 3.56 | 0.009 |
| Polyuria | Absent | 2.08 | 1.27, 3.43 | 0.004 |
| Abdominal circumference code | Normal | 0.48 | 0.33, 0.71 | 0.0002 |
| Occupation | Working |  |  |  |
| Housewife |  | 0.65 | 0.42, 1.00 | 0.009 |
| Not working |  | 1.58 |  | 0.033 |

B. Continuous variables
| Variable | Beta | SE | P-value |
| --- | --- | --- | --- |
| Age | 0.021 | 0.010 | 0.047 |
| Duration of DM | 0.102 | 0.016 | <0.0001 |
| Fasting Plasma Glucose | 0.003 | 0.001 | 0.009 |
| Abdominal Circumference | 0.017 | 0.007 | 0.018 |
| Duration of HTN | 0.068 | 0.018 | 0.0002 |

Stepwise regression identified eight variables for inclusion in the model: polyuria, ischemic heart disease, stroke, neuropathy, abdominal circumference code, fasting plasma glucose, systolic blood pressure, and duration of DM. In the final stepwise analysis polyuria, ischemic heart disease, stroke, abdominal circumference code, fasting plasma glucose and duration of DM all associated with DR (**Table 8**). Polyuria and stroke both associated with increased odds of DR, while presence of ischemic heart disease and obesity as measured by abdominal circumference code associated with reduced risk. Both higher fasting plasma glucose and longer duration of DM increased risk significantly (**Table 8**).

**Table 8.** Hypertension only – Stepwise multivariate regression.

| **Variable** | **Referent** | **OR** | **95% CI** | **P-value** |
| --- | --- | --- | --- | --- |
| Polyuria | No | 1.99 | 1.12, 3.54 | 0.020 |
| Ischemic Heart Disease | No | 0.38 | 0.19, 0.78 | 0.008 |
| Stroke | No | 2.68 | 1.00, 7.15 | 0.049 |
| Neuropathy | No | 2.27 | 0.92, 5.62 | 0.077 |
| Abdominal Circumference Code | Normal | 0.51 | 0.33, 0.77 | 0.001 |
|  |  | Beta | SE |  |
| Fasting Plasma Glucose |  | 0.003 | 0.001 | 0.030 |
| Systolic Blood Pressure |  | 0.005 | 0.004 | 0.246 |
| Duration of DM |  | 0.100 | 0.017 | <0.0001 |

### Mediation analyses

As there is a possibility that the effect of duration of diabetes on DR is affected by the age of onset, we performed a mediation analyses to test this hypothesis(36). We used the mediator model (with age of onset (mediator) as the dependent variable and duration of diabetes as the independent variable) with an outcome of DR. We performed an unadjusted model and found that 8.6% of the effect of duration of DM was mediated by the age of onset (p = 0.012). The effect of age of onset represents a significant causal contribution to DR. However, a minority of the risk due to duration of DM was mediated by age of onset. Hence both variables were necessary to fully model DR risk. This may explain why under some models age of onset is significant, but in others it is not.

## Discussion

Our study of an Indian slum population at high risk of DR revealed several clear patterns of association. The most consistent factor in terms of association across all our analyses was duration of DM; this result was partially mediated by age of onset. Individuals with longer duration and earlier onset of DM were at increased risk of developing DR. This result was consistent with the prior literature in numerous populations (37-43). However, our sample had the advantage of being population based as opposed to hospital or clinic based, which is more the norm (41). Another factor that is consistent with prior literature from diverse populations was the association of DR with poorer glycaemic control as measured by either FPG or HbA1C (41, 44-47). Poorer control increased risk of DR.

We also found that increasing BMI and other measures of obesity associated with protection from DR. This pattern of association is nowhere near universal in the literature. In several studies from multiple countries, there was no association between DR and measures of obesity(18, 41, 48, 49). Other studies have shown that increasing BMI or obesity measured using various metrics increased risk, especially among those who are obese, and, in some cases, obesity associated with more severe DR (PDR vs NPDR) (50-54). However, in studies that had results showing positive association between obesity and DR the average BMI was usually higher than in our study population with sample means over 30 as compared to mean values ∼26 in our Mumbai slum sample. Other studies in Asia found patterns similar to ours with higher levels of obesity being protective and average BMIs being lower (55, 56). However, even in Asians the relationships may be complex with some of the associations being only in females and in these studies obesity increased risk (57). The complexity of the relationship between obesity and DR is further exemplified by one study in the US where it was found that being either underweight or obese associates with DR compared to people of normal BMIs in univariate analyses, but only being underweight associated with increased risk in a multivariable analysis (58). Again, in these studies baseline BMI was greater than in our study. Finally, in some cases, the differences may be attributable to T1 vs T2DM. For example, among T1DM, measures of central of obesity positively associated with DR in a Finnish study (51). As we did not explicitly assess T1DM versus T2DM in our sample this may affect results. However, as we note above the likelihood of having many T1DM cases in our sample is low.

Similar to our results, a prior study in India showed that high BMI was protective, although isolated abdominal obesity associated with increased DR risk (57). Another study in Singapore also showed protection with increasing BMI, but WHR positively associated with DR and DR severity in women (56). Our results were more uniform in the pattern of association with DR in that all associations between obesity measures and DR showed increasing obesity associated with protection. In the prior Indian study average BMI was low, 24.4 for men and 26.5 for women, and in the Singapore study the average BMI was <26 for all participants. Another possibility is that the type of DR differs between our population and that reported in prior studies. For example, we found ∼8% of the DR subjects had PDR and this may be higher than other studies.

A prior meta-analysis of 13 diverse prospective cohort studies showed association of obesity with DR overall but not with PDR as a distinct outcome, but this analysis did not account for baseline levels of obesity among the populations nor the ancestry of the participants (49). However, in this study subgroup analyses failed to reveal significance, indicating a complex pattern of associations among studies. Overall, it would appear that the pattern of association differs by DR subtype and between Asians and Europeans, although there is some variability even within continents. Factors that underlie these differences are unclear but as pointed out may relate to differences in population average BMI or in genetic or environmental differences that were unmeasured.

We also identified not only different risk in men and women with men being at higher risk, but some different patterns of association by sex. This is consistent with prior work that elucidated variation in DM sequalae by sex (59).

Systolic blood pressure also associated positively with DR, but in sex stratified analyses SBP was only associated in males, indicating blood pressure may act differently in the two sexes. This may be due to BP differences by sex as men tend to have higher BP in India (60). For example, in a sample of almost 9000 people, males in rural India have higher prevalence of prehypertension and hypertension but the transition from prehypertension to hypertension over a five-year window was more common among females (61). Our multivariate analysis of only hypertensives showed no association with SBP which was unsurprising. However, unlike in prior analyses with all participants where age negatively associated with DR, age positively associated with DR among hypertensives. This may be related to the fact that hypertensives are in general older than the general population as was the case for our cohort.

Our study provided strong evidence that duration of DM was strongly associated with DR. This is completely consistent with prior reports in the literature. We were able to show, however, that a significant albeit minority part of this association was due to age of DM diagnosis. In contrast to the consistency with the literature with respect to DM duration, our results for measures of obesity exemplify the complexity of the relationship between body mass measures and DR. Our findings indicate that all measures of obesity negatively associated with DR. This may in part be due to the nature of our study population that had an overall BMI lower than most other studies. However, even compared to other low BMI Asian samples, our study was not completely consistent because even in studies where BMI patterns were similar to ours measures of central obesity associated differently and in sex specific ways. Our results highlight both the complex nature of the DR associations and diverse patterns among populations.

## Data Availability

Data will be deposited in Dryad and openly available upon acceptance.

## Author Contributions

Substantial contributions to conception and design, acquisition of data or analysis and interpretation of data: R.K., A.J., S.N, R. S., J.B., S.K.I., S.M.W., and N.S.

Drafting the article or revising it critically for important intellectual content: : R.K., A.J., S.N, R. S., J.B., S.K.I., S.M.W., and N.S.

Final approval of the version to be published: : R.K., A.J., S.N, R. S., J.B., S.K.I., S.M.W., and N.S.

